# Tolerability and Pharmacokinetic Evaluation of Inhaled Dry Powder Hydroxychloroquine in Healthy Volunteers

**DOI:** 10.1101/2020.12.03.20243162

**Authors:** Y.A. de Reus, P. Hagedoorn, M.G.G. Sturkenboom, F. Grasmeijer, M.S. Bolhuis, I. Sibum, H.A.M. Kerstjens, H.W. Frijlink, O.W. Akkerman

**Affiliations:** University of Groningen, University Medical Center Groningen, Department of Pulmonary Diseases and Tuberculosis, Groningen, the Netherlands; University of Groningen, Department of Pharmaceutical Technology and Biopharmacy, Groningen, the Netherlands; University of Groningen, University Medical Center Groningen, Groningen, Department of Clinical Pharmacy and Pharmacology, the Netherlands; PureIMS B.V., Roden, the Netherlands; University of Groningen, University Medical Center Groningen, TB center Beatrixoord, Groningen, the Netherlands

**Author notes:** Corresponding author Onno W. Akkerman, MD, PhD, University Medical Center Groningen, PO Box 30001, 9700 RB Groningen, the Netherlands.

**Keywords:** SARS-CoV-2, covid-19, hydroxychloroquine, inhalation

## Abstract

**Rationale:** Inhaled antimicrobials enable high local concentrations where needed and, compared to orally administration, greatly reduce the potential for systemic side effects. In SARS-CoV-2 infections, hydroxychloroquine (HCQ) administered as dry powder via inhalation could be safer than oral HCQ allowing for higher and therefore more effective pulmonary concentrations without dose limiting toxic effects.

**Objectives:** To assess the local tolerability, safety and pharmacokinetic parameters of HCQ inhalations in single ascending doses of 5, 10 and 20 mg using the Cyclops dry powder inhaler.

**Methods:** 12healthy volunteers were trained in inhaling HCQ correctly. Local tolerability and safety were assessed by pulmonary function tests, ECG and recording adverse events. To estimate systemic exposure, serum samples were collected before and 0.5, 2 and 3.5 h after inhalation.

**Results and discussion:** Dry powder HCQ inhalations were well tolerated by the participants, except for transient bitter taste in all participants and minor coughing irritation. There was no significant change in QTc-interval or drop in FEV_1_ post inhalation. The serum HCQ concentration remained below 10 µg/L in all samples.

**Conclusion:** Inhaled dry powder HCQ is safe and well tolerated. Our data support further studies with inhaled HCQ dry powder to evaluate pulmonary pharmacokinetics and efficacy is warranted.

## 1. INTRODUCTION

In late December 2019 an outbreak of the novel coronavirus, Severe Acute Respiratory Syndrome Coronavirus 2 (SARS-CoV-2), started in Wuhan, China, and caused the spread of corona virus disease 2019 (COVID-19) [1]. The WHO declared the epidemic of COVID-19 a pandemic on March 12th, 2020 [2]. The virus is still rapidly spreading with over 41 million cases and 1.1 million deaths reported worldwide in the second week of November 2020 [3]. Currently Europe is again facing a steep rise in incidence and is in its ‘second wave’ now autumn has come.

Given that this pandemic has a huge impact on healthcare systems, social life and economics, there is an urgent need for treatment of COVID-19. Currently available treatment options are mostly used in hospitalized patient: the anti-inflammatory drug dexamethasone and anti-viral nucleoside analog prodrug remdesivir. These drugs reduce mortality and shorten time to recovery respectively [4, 5]. In addition to an effective treatment there is also the need to be able to prevent and decrease transmission in the general population and moreover in healthcare workers or other high-risk groups. We are awaiting results from clinical trials with vaccines; as these will almost certainly not be 100% effective, alternatives should be investigated for both treatment in early disease and prevention of transmission. Repositioning old drugs for use as antiviral or anti-inflammatory treatment is an interesting strategy because the safety profile, side effects, posology and drug interactions are already known which can speed up the trial program duration considerably.

Among those drugs is hydroxychloroquine sulphate (HCQ), which is mostly used in rheumatologic conditions because of its immune-modulatory effects [6, 7]. HCQ has proven to be effective in in vitro Vero cell systems infected with SARS-CoV-2 in two separate Chinese studies [8, 9]. Angiotensin converting enzyme 2 (ACE2) receptor expressing cells play an important role in the pathogenesis of SARS-CoV-2 infection, as the virus uses this receptor for entering the cell [8, 10-12]. HCQ impairs the terminal glycosylation of ACE2 and thereby inhibits cell-binding and consequently entry of the virus into the cell [13-15]. Furthermore, HCQ also blocks transport of SARS-CoV-2 from early endosomes to endolysosomes, a requirement to release the viral genome [8, 9, 16]. Finally, HCQ has anti-inflammatory properties as it influences the generation of pro-inflammatory cytokines and endosomal inhibition of toll-like receptors, which have a major role in innate immune response [8, 17-20]. Based on these in vitro findings oral HCQ was used abundantly worldwide in the beginning of the COVID-19 pandemic, both off label and in clinical trials. Some observational studies showed clinical benefit and antiviral effects [21-25] while others did not [26, 27] or were inconclusive [28, 29]. Currently both the FDA and EMA advice against the off-label use of oral HCQ based on the large clinical RECOVERY trial that showed no beneficial effects on 28-day mortality [30]. The prospective European DISCOVERY trial and WHO SOLIDARITY trial have discontinued the oral HCQ treatment because of a lack of effect on mortality arms as well. The failing treatment with oral HCQ may be explained by insufficient concentrations in alveolar epithelial cells due to its enormous volume of distribution of 5500 L [31]. Raising the oral dose is not an option since this is limited by adverse or even toxic effects, including the risk of cardiovascular toxicity (QTc prolongation).

Pulmonary administration of HCQ can be the solution to reach high local pulmonary concentrations without systemic toxicity [32, 33]. For this purpose, we developed a dry powder formulation of HCQ suitable for inhalation using the Cyclops dry powder inhaler. The Cyclops is a high dose disposable inhaler that enables effective dispersion of up to 50 mg of drug in the preferred size range for inhalation [34]. The aim of this study was to assess local tolerability and safety of increasing doses of dry powder HCQ administered using the Cyclops to healthy volunteers. The chosen doses of 5 mg, 10 mg and 20 mg are comparable with known doses of inhaled nebulized HCQ and thus expected to be safe [33, 35-38]

## 2. METHODS

### Study design

This study was an open-label phase 1a single ascending dose study with twelve healthy volunteers. It was performed at the University Medical Center Groningen (UMCG) location Beatrixoord (Groningen, the Netherlands) between in September and October 2020. It included administration of HCQ dry powder per inhalation using the Cyclops in single ascending doses of 5 mg, 10 mg, and 20 mg. In- and exclusion criteria are listed in Table 1. The study was approved by the local medical ethical review committee (METc UMCG, Groningen, The Netherlands, METc number 2020.168). The study was performed according to the Helsinki declaration (Fortaleza, Brazil, 2013) and was registered at clinicaltrials.gov (NCT04497519). The study was conducted with the written informed consent of all participating subjects.

**Table 1.**
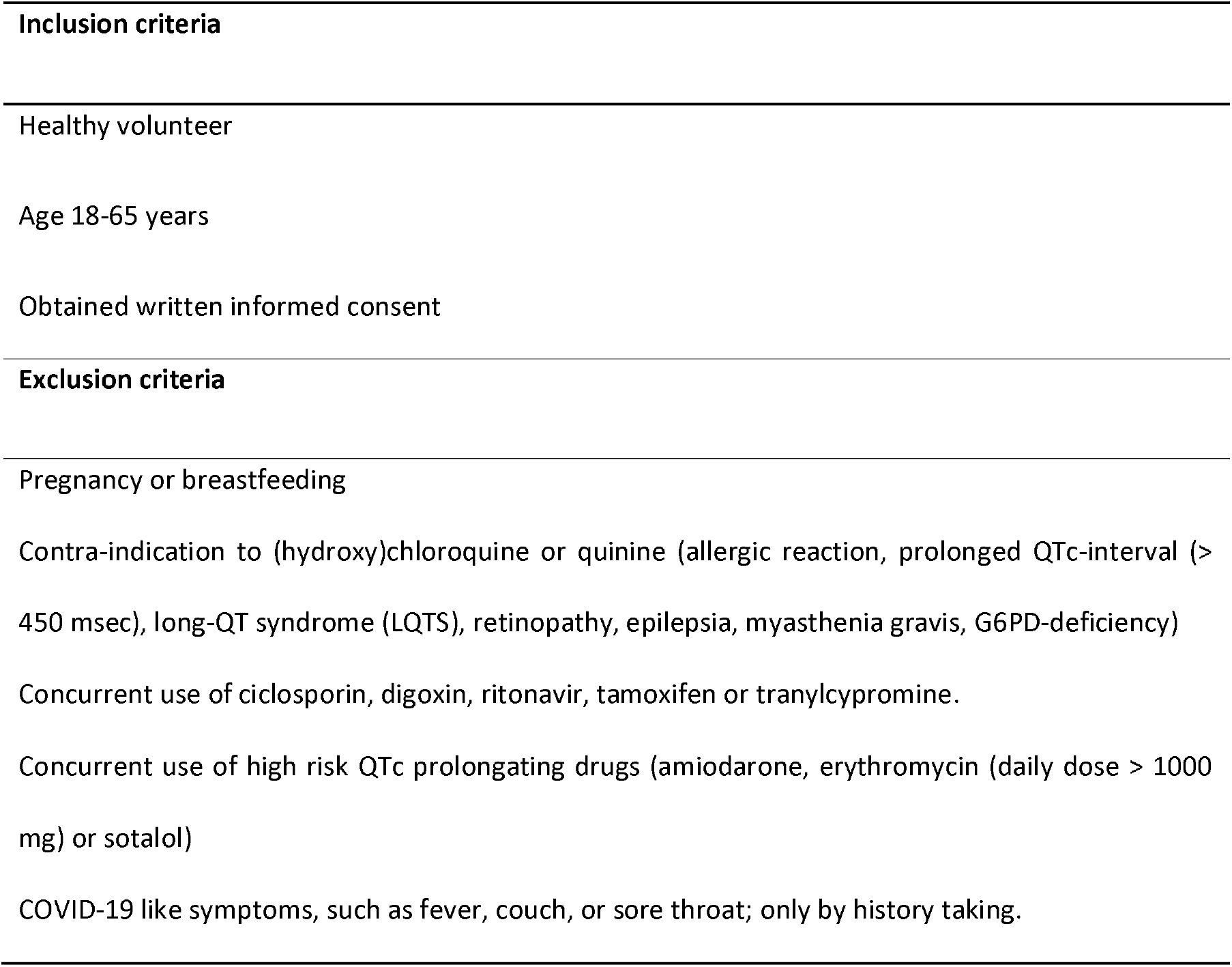
In- and exclusion criteria

### Study drug

The dry powder formulation of HCQ was developed at the department of Pharmaceutical Technology and Biopharmacy of the University of Groningen. For the production of the study drug hydroxychloroquine sulphate was obtained from Ofipharma BV (Ter Apel, the Netherlands). The HCQ Cyclops was produced by PureIMS B.V. (Roden, the Netherlands). Each Cyclops contained a nominal dose of 5 mg or 10 mg HCQ; the 20 mg dose was administered as 2 successive inhalations with 10 mg of HCQ.

### Objectives and procedures

The primary objective was to assess local tolerability and safety. Local tolerability was assessed by spirometry combined with active questioning about adverse events experienced by the participants. A drop of the forced expiratory volume in the first second (FEV_1_) of 15% or more after inhalation of HCQ compared to baseline FEV_1_ was considered clinically significant and critical to decide on proceeding with the next ascending dose. Spirometry was performed before inhalation (baseline) and 35 minutes and 95 minutes after inhalation of HCQ according to the ATS/ERS guidelines [39]. Adverse events were continually assessed during the study day. Cough for more than one hour or any other reported adverse event that made either the physician or the participant decide to stop participation was considered critical to decide on proceeding with the next ascending dose. ECGs were performed to assess the QTc interval as safety parameter. An ECG was obtained at the screening visit, before inhalation of the first dose and approximately 3.5 hours after each HCQ inhalation. An observed QTc interval of more than 500 ms was also considered critical on proceeding with the next ascending dose. All tolerability and safety endpoints were discussed with a Data Safety Monitoring Board (DSMB) after all twelve participants completed a dose step before proceeding to the next ascending dose.

The secondary objectives were to assess systemic exposure of HCQ and measurement of inspiratory flow parameters. To determine the systemic exposure, blood samples were collected from an intravenous indwelling cannula just before inhalation and 30 minutes, 2 hours and 3.5 hours after inhalation of HCQ. These samples were analyzed using a validated liquid chromatography tandem mass spectrometry (LC-MS/MS) method performed at the laboratory of the department of Clinical Pharmacy and Pharmacology of the UMC Groningen (ISO15189:2012 (M170) certified). The limit of quantification is 10 µg/L. The delivered dose was determined by subtracting the powder residue inside the Cyclops after inhalation from the exact weighed dose pre inhalation. The powder residues in the Cyclops were dissolved in demineralized water and the solutions were analyzed with a Thermo Scientific spectrophotometer (Genesys 150 UV–VIS, The Netherlands) at a wavelength of 236.0 nm.

Prior to inhalation of the study drug, study participants received inhalation instructions followed by training regarding handling of the device and performing a correct inhalation maneuver. Training was done using an empty Cyclops connected to a laptop, with in-house developed software application (labVIEW, National Instruments, the Netherlands) for recording of and processing of flow curves generated through the device. When a series of consistent flow curves meeting the criteria for good inhaler performance was obtained during training, a similarly instrumented Cyclops with HCQ was handed to the participant. Inspiratory flow parameters were recorded during each drug administration.

### Data management and statistics

Study data were collected and managed using Research Electronic Data Capture (REDCap) [40, 41]. Statistical analysis was performed with SPSS version 23. Data from all eligible subjects who received at least one dose of the study drug were included in analyses of safety. Data were summarized using descriptive statistics. At each visit and timepoint, testing for differences in pre- to post-dose changes in FEV1 (liters) and QTc interval were performed using the paired T-test or Wilcoxon rank sum test. P values below 0.05 were considered statistically significant.

## RESULTS

Twelve healthy volunteers were enrolled, and all completed the study. See table 2 for all patient characteristics. All participants had normal baseline QTc interval and G6PD-deficiency was excluded in all participants. At the screenings visit pregnancy was excluded in female participants.

**Table 2.**
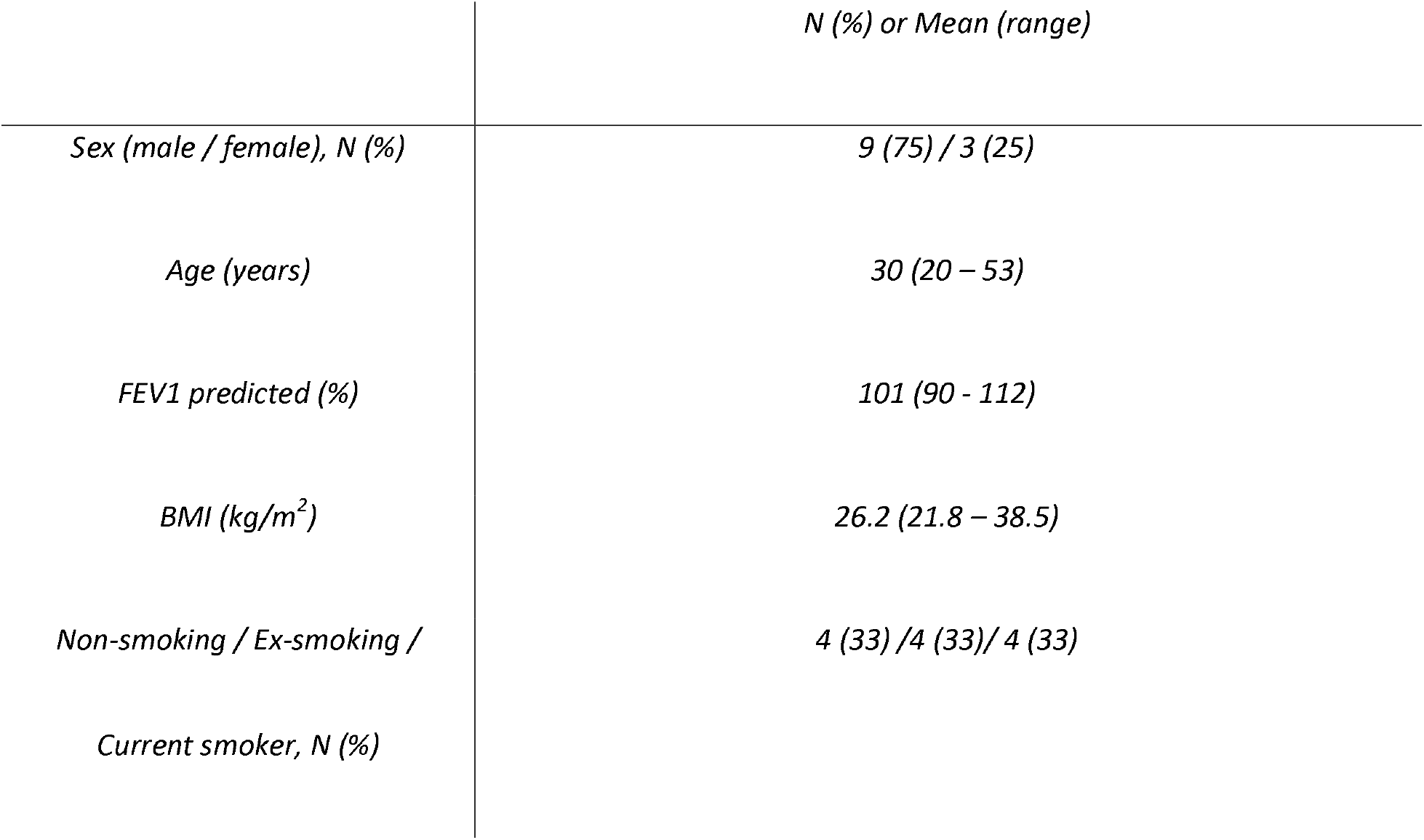
Participant characteristics

There were no serious adverse events observed during or after the study (see table 3). None of the participants had cough longer than the pre-defined safety period of one hour. Minor complaints of cough were reported four times, each in a different participant, directly after inhalation of HCQ; two times after a dose of 10 mg HCQ and two times after a dose of 20 mg HCQ. Complaints varied from experience of an itchy or tickling sensation in the throat to a single or a few observed coughs that were self-limiting. Two participants reported minimal dyspnea which disappeared after coughing or spontaneously within 4 minutes after inhalation. All participants mentioned bitter taste after inhalation. In most participants this lasted for 5-10 minutes, but one participant reported the bitter taste for 2 hours. Participants were advised to rinse their mouth with water or eat something after the dose was administered with satisfying effect. Two participants had complaints of slight nausea relating to the bitter taste; one for 5 minutes after a dose of 5 mg HCQ and one for 70 minutes after a dose of 10 mg HCQ. Sore throat was reported by three participants. Two participants developed symptoms of upper respiratory tract infection in the following days of which one tested positive by PCR nasopharyngeal swab for SARS-CoV-2 two days after the last study visit. One participant experienced some hoarseness, which disappeared after drinking some water. Two other adverse events were recorded. One participant with an intrauterine contraceptive device in situ experienced minor spotting two days after inhalation of HCQ after both the 5 mg and 10 mg dose. This is not mentioned as a known side-effect of oral HCQ [42]. One participant mentioned dry eyes once at a dose of 20 mg, which seemed to be related to the dry hospital environment and not to administration of the study drug.

**Table 3.** Reported adverse events out of 36 HCQ administrations by inhalation

| **Adverse events** | **Percentage of total administrations** | **Percentage of patients** |
| --- | --- | --- |
| Cough | 11.1% | 33.3% |
| Dyspnea | 5.6% | 16.7% |
| Bitter taste | 100% | 100% |
| Nausea | 5.6% | 16.7% |
| Sore throat /<br>respiratory infection | 8.3% | 25% |
| Hoarseness | 2.8% | 8.3% |
| Spotting | 5.6% | 8.3% |
| Dry eyes | 2.8% | 8.3% |

None of the participants showed a significant drop in FEV_1_ (≥15%) at any time point after HCQ administration (see table 4). A maximum drop of -7.51% was observed 95 minutes after inhalation of 5 mg HCQ. This participant mentioned a sort of burning sensation at the chest at that moment as well but no dyspnea, something he also recognizes while running.

**Table 4.**
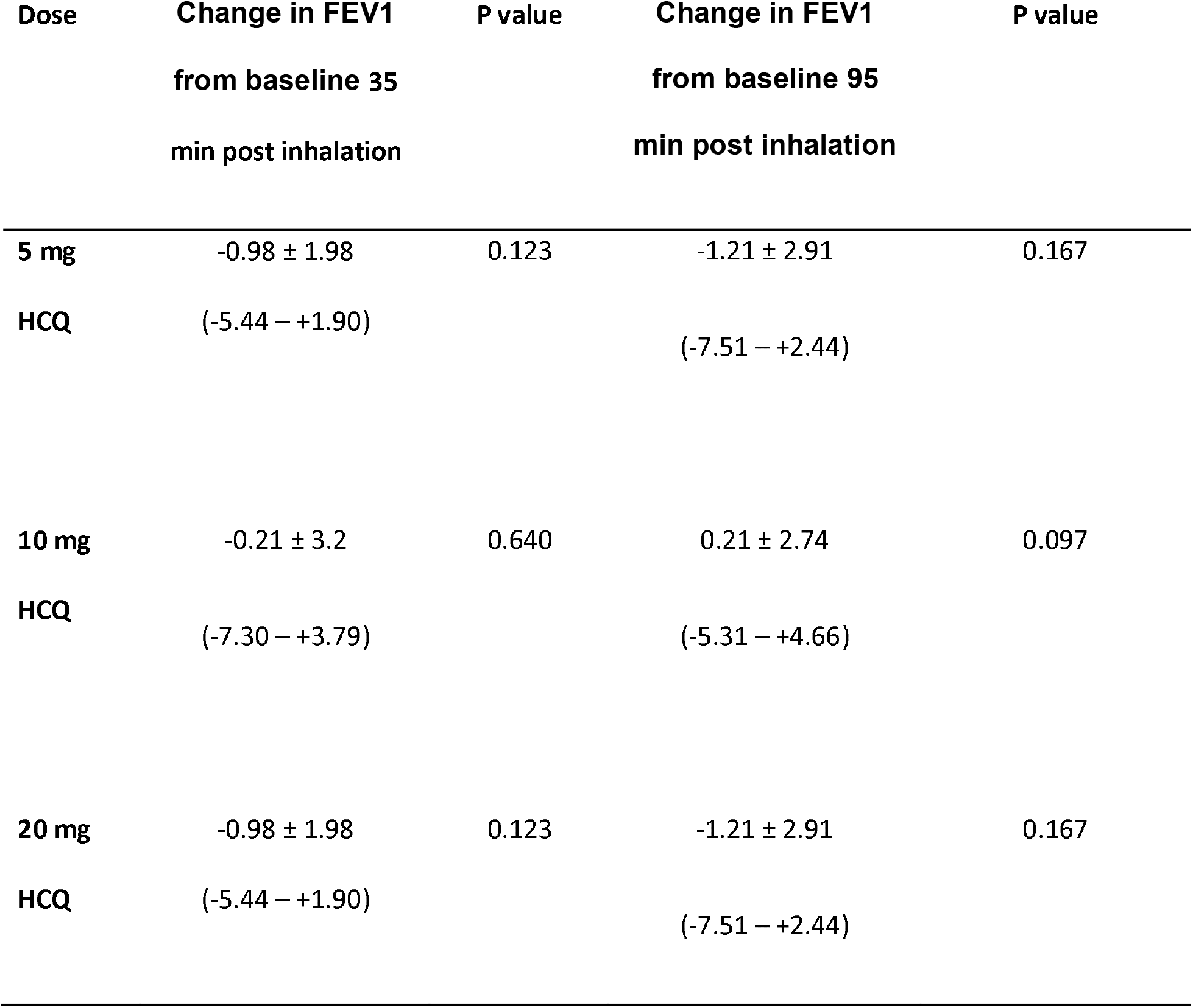
change in FEV1 post inhalation compared to baseline in %: mean ± SD and range

None of the participants had a QTc prolongation ≥ 500, nor even ≥ 450 ms. Mean QTc interval was 412 ± 21 ms (range 384-441) at baseline. This did not change significantly 3.5 hour after inhalation of 5 mg, 10 mg and 20 mg HCQ, respectively (see table 5).

**Table 5.** Change in QTc time in ms

|  | 3,5 h after<br>administration | P value |
| --- | --- | --- |
| Before administration | 412 ± 21 ms<br>(range 384-441) |  |
| Post 5 mg HCQ | 407 ± 19 ms (range<br>383-439) | 0.538 |
| Post 10 mg HCQ | 414 ± 15 ms<br>(range 392-447) | 0.736 |
| Post 20 mg HCQ | 409 ± 12 ms (range<br>381-423) | 0.648 |

In all participants, the serum HCQ concentrations sampled 30 minutes, 2 hours and 3.5 hours, were below the detection level of 10 µg/L irrespective of dose. The mean delivered dose was 3.16 mg (63%) after administration of 5 mg HCQ, 6.95 mg (70%) after administration of 10 mg HCQ and 14.86 mg (75%) after administration of 20 mg HCQ. Based on the recorded inspiratory flow parameters, all participants correctly performed the inhalation maneuvers.

## DISCUSSION

Pulmonary administration of HCQ for early COVID-19 treatment or prevention in post-exposed individuals might be better than oral administration of HCQ due to the pharmacological properties of HCQ. Hypothetically, it can reach higher pulmonary concentrations compared to oral HCQ while using much lower doses and exerting lower systemic exposure. This phase 1 study with inhaled HCQ with three different doses showed good local tolerability and safety without significant systemic side effects.

Coughing was reported four times out of a total of 36 administered doses (11%) in four different participants and was very mild in severity. Coughing is often reported after inhalation of antimicrobials by both wet nebulization and dry powder inhalation, with data mainly available from cystic fibrosis patients treated with colistin or tobramycin. In general, this population cough is more frequently reported after dry powder inhalation (ranging from 75% - 90%) than after nebulization (ranging from 31% - 78%) in these patients, although some studies found no differences [43-45]. The trigger for both cough and probably bitter taste is deposition of the drug in the oropharynx. This could be expected since HCQ is a quinoline known for its extremely bitter taste (249 on a bitter scale compared to caffeine at 46) [46]. However, the majority reported that the taste was not disturbing since the participants were warned beforehand, it was minor, and it disappeared within a few minutes after HCQ inhalation or even faster when rinsing the mouth with water or eating something directly after inhalation.

Bronchus obstruction was not a problem after inhalation of HCQ dry powder; in none of the twelve participants experienced a drop in FEV_1_ of more than 15%. The maximum drop was 7.5 % compared to baseline and this was not accompanied by dyspnea. One participant did mention a light burning sensation at the chest at that moment, something he also experienced while running. Although not formerly diagnosed, this participant might suffer from a mild exercise induced asthma. Nevertheless, we believe that HCQ dry powder can be used safely in asthma patients as well, since aerosolized HCQ has been applied in a phase I clinical study to assess safety for use in asthma and was concluded to be safe and well tolerated in 31 healthy individuals in doses up to 20 mg daily for 7 days [33, 36]. In 2006, a phase 2 clinical trial with aerosolized HCQ as anti-inflammatory treatment for patients with asthma followed. A dose of 20 mg daily was tolerated for up to 21 days, but it failed to meet the primary clinical endpoints for effective asthma treatment; relative improvement in FEV_1_ compared to baseline was not statistically significant after treatment compared to placebo [33, 37]. None of these participants had significant ECG changes and side effects consisted of headache and nausea only [33, 36, 37].

HCQ serum concentrations were below the quantification limit of 10 µg/L in all participants and irrespective of dose or timepoints. Possibly our timing of blood sampling was sub optimal. The timepoints were chosen based on the absorption rate after oral administration, because data after inhalation were not available at that time. If the maximum concentration (C_max_) occurs very shortly after inhalation (T_max_), we might have missed this peak concentration with our first blood sample drawn after 30 minutes. This is supported by the only other available pharmacokinetic data from a phase I clinical trial with aerosolized HCQ that came available after our study protocol was developed. Fifteen healthy volunteers inhaled single doses of 5, 10 or 20 mg HCQ. Reported HCQ serum concentrations were mean C_max_ between 22 and 69 µg/L with an early T_max_ within 2-3 minutes. The reported systemic exposure was very low (7-54 µg*h/L), suggesting elimination within 30 minutes [33].

Local lung concentrations are expected to be higher after inhalation of HCQ compared to oral administration, even though the highest dose of inhaled HCQ in this study (20 mg) is only a fraction of the usual oral HCQ dose (200-800 mg). For example, if the delivered dose of 14.86 mg homogeneously distributes over the lung tissue (843 mL) [47], then a lung tissue HCQ concentration of approximately 40 µM would be achieved. Preliminary results from our own experiments in primary human epithelial cells indicate that HCQ concentrations of approximately 20 to 40 µM do result in a significant reduction in viral load after SARS-CoV-2 infection (manuscript in preparation). This is in contrast to the lack of antiviral effect found by Mulay et al. at an HCQ concentration of only 10 µM, which because of the fourfold lower concentration than potentially achievable after HCQ inhalation seems to hold less relevance [48]. Based on these considerations, effective concentrations can potentially be achieved in lung tissue after inhalation of 20 mg HCQ. These high HCQ concentrations and the superior potential of inhaled over oral HCQ should be the new starting point of any further clinical studies on the activity of HCQ against SARS-CoV-2.

The concerns about cardiotoxicity of oral HCQ, a well-known drug and generally considered to be safe and well tolerated, were magnified by a large retrospective observational study that reported a strong association between the use of HCQ and ventricular tachycardia and death in hospitalized COVID-19 patients [50]. However, this paper was retracted soon after publication because of concerns about the data validity [49, 50]. White *et al*. state that a lot of confusion has arisen by extrapolating long-term risks of myocardial damage with chronic dosing to short-term exposures, thereby overestimating the risk of ventricular arrhythmias [31]. Our study shows that systemic exposure after inhalation is very low, which is a positive result regarding the risk of systemic toxicity. Also, the concerns for any other possible systemic adverse event should be tempered because of these results.

A limitation of our study is that only serum concentrations are measured and not local pulmonary concentrations. So far, only in vitro experiments on Vero cells have shown efficacy and data from human pulmonary concentrations and thus local efficacy are lacking [8, 9]. Since in our study no systemic exposure of HCQ was detected for all participants and for all doses, it is not expected that systemic side effects will occur and HCQ might therefore also be used in patients of older age and with comorbidities.

In conclusion, HCQ inhalation using the Cyclops is safe and generally well tolerated by healthy volunteers, except for minor cough and bitter taste. These positive results and the superior safety and efficacy potential of inhaled over oral HCQ strongly encourage the execution of further clinical studies with inhaled HCQ to battle this COVID-19 pandemic.

## Data Availability

Data is available, however some restrictions will apply. Requests can be sent to the ethical board

## Acknowledgements

We thank Marieke van Rossum and Marina Zenina for their help in the execution of this study.

## Funding Sources

The production, labelling and distribution of the HCQ dry powder Cyclops inhalers was provided free of charge by PureIMS.

All other funding was supplied by the institutions of the participating authors at University of Groningen and University Medical Center Groningen

## Conflict of interest

**YdR**: no conflicts of interest

**PH**: The employer of PH has a royalty agreement with AstraZeneca on the sales of the Genuair Inhaler; In addition, PH has a patent WO2015NL50413 20150605 licensed.

**MS**: no conflict of interest

**FG**: FG is employed by PureIMS, the manufacturer of the Cyclops inhaler.

**MB**: no conflicts of interest

**IS**: no conflicts of interest HK: no conflict of interest

**EF**: The employer of HWF has a license agreement with PureIMS on the Cyclops patent (WO2015/187025)

**OA**: no conflicts of interest

## Notes

### Clinical Trial

NCT04497519

### Author Declarations

The study was approved by the local medical ethical review committee (METc UMCG, Groningen, The Netherlands, METc number 2020.168).

